## Supplementary material for "Leptospirosis in the Caribbean Region between 2000 and 2022 – a Scoping Review of Morbidity and Mortality": S2 Table

**Supporting Table 2. Full description of the search strategy used in each database and the number of publications retrieved.**

| **Database and search strategy** | **Results** |
| --- | --- |
| **PubMed** | **505** |
| ("Leptospirosis"[Mesh] and Leptospirosis [tiab] or Leptospiroses [tiab] or "Leptospira Infection" [tiab] or "Infection, Leptospira" [tiab] or "NEGLECTED DISEASES" or "Neglected tropical diseases" [tiab] or "zoonosis" [tiab] or "Leptospirosis Canicola" [tiab] or zoonotic [tiab] or "Leptospirosis" [tiab])  AND  ("Caribbean Region"[Mesh] OR "Caribbean Region" [tiab]  OR Haiti [MESH] or Haitian OR "Dominican Republic" [MESH] OR Dominicans OR Cuba [MESH] OR Cuban OR "Puerto Rico" MESH] OR "Puerto Ricans" OR Jamaica [MESH] OR Jamaicans OR "Trinidad and Tobago" [MESH] OR "Trinidad* and Tobago*" OR Guyana [MESH] OR Guyanese OR Suriname [MESH] OR Suriname* OR Bahamas [MESH] OR Bahamians Belize [MESH] OR Belizeans OR Guadeloupe [MESH] OR Guadeloupeans OR Martinique [MESH] OR Martinicans OR Barbados [MESH] OR Barbadians OR Curacao [MESH] OR Curaçaoans OR Curaçao [MESH] OR Curaçaoans OR "Saint Lucia" [MESH] OR Saint Lucians OR Grenada [MESH] or Grenadians OR Aruba [MESH] OR Arubans OR "Saint Vincent and the Grenadines" [MESH] OR "Vincentians*" OR U.S. Virgin Islanders OR "Antigua and Barbuda" [MESH] OR "Antigua* and Barbuda*" OR Dominica [MESH] OR Dominicans OR "Cayman Islands" OR "Saint Kitts and Nevis" [MESH] or "Saint Kitts* and Nevis*" OR "Turks* and Caicos* Islands*" OR "Sint Maarten" [MESH] OR "Sint Maarten*" OR "Saint Martin*" OR "British Virgin Islands" [MESH] OR "British* Virgin* Islands*" OR "Caribbean Netherlands" [MESH] OR "Caribbean* Netherlands*" OR Anguilla [MESH] OR Anguilla* OR "Saint Barthélemy*" OR Barthelemy* OR St. Barts* OR Montserrat [MESH] OR Montserratian OR "Caribbean countries") OR "French Guiana" [MeSH] OR "French* Guiana*" OR "French Guiana" [tiab] | |
| **Web of Science** | **592** |
| (TS=(Leptospirosis) and TI=(leptospirosis) and AB = (leptospirosis) or TS = (Leptospira)  AND AB = (Leptospira) and AB =("Neglected Tropical Disease") and TI = (Neglected Tropical Disease) and TI = (zoonosis) and AB=(zoonosis) or TS = ("Leptospirosis Canicola") and TI = (zoonotic) and AB = (zoonotic) and TI = ("Leptospirosis Canicola ") and AB = ("Leptospirosis Canicola") AND TS=(Caribbean) and TI=(Caribbean) and AB = (Caribbean or Haiti OR "Dominican Republic" OR Cuba OR "Puerto Rico" OR Jamaica OR "Trinidad and Tobago" OR Guyana OR Suriname OR Bahamas OR Belize OR Guadeloupe OR Martinique OR Barbados OR Curacao OR Curaçao OR "Saint Lucia" OR Grenada OR Aruba OR "Saint Vincent and the Grenadines" OR "United States Virgin Islands" OR "Antigua and Barbuda" OR Dominica OR "Cayman Islands" OR "Saint Kitts and Nevis" OR "Turks and Caicos Islands" OR "Sint Maarten" OR "Saint Martin" OR "British Virgin Islands" OR "Caribbean Netherlands" OR Anguilla OR "Saint Barthélemy" OR "Saint Barthelemy" OR “Montserrat” or “British Guiana” or “French West Indies” or “British West Indies”) AND SU=(human)) AND (DT==("ARTICLE")) | |
| **Scopus** | **309** |
| ALL ( leptospirosis ) OR ABS ( zoonosis ) OR KEY ( zoonosis ) OR KEY ( "neglected tropical disease" ) AND ALL ( "leptospira infection" ) AND ( ALL ( Caribbean ) OR ALL ( human ) OR ALL ( prevalence ) OR ALL ( incidence ) OR ALL ( "Caribbean countries" ) ) AND ( Haiti OR "Dominican Republic" OR Cuba OR "Puerto Rico" OR Jamaica OR "Trinidad and Tobago" OR Guyana OR Suriname OR Bahamas OR Belize OR Guadeloupe OR Martinique OR Barbados OR curacao OR cura&#231;ao OR "Saint Lucia" OR Grenada OR Aruba OR "Saint Vincent and the Grenadines" OR "United States Virgin Islands" OR "Antigua and Barbuda" OR Dominica OR "Cayman Islands" OR "Saint Kitts and Nevis" OR "Turks and Caicos Islands" OR "Sint Maarten" OR "Saint Martin" OR "British Virgin Islands" OR "Caribbean Netherlands" OR Anguilla OR "Saint Barth&#233;lemy" OR "Saint Barthelemy" OR Montserrat OR TITLE-ABS-KEY ( "British Guiana" ) OR TITLE-ABS-KEY ( "British West Indies" ) OR TITLE-ABS-KEY ( "French West Indies" ) ) AND ( LIMIT-TO ( DOCTYPE , "ar" ) ) | |
| **Embase** | **323** |
| ('leptospirosis' OR leptospiroses:ab,ti OR 'leptospira infection':ab,ti OR 'neglected disease':kw OR 'neglected tropical disease':ab,ti OR 'zoonosis':ab OR ((((((Haiti:ab OR 'Dominican Republic':ab OR Cuba:ab OR 'Puerto Rico':ab OR Jamaica:ab OR Trinidad:ab) AND Tobago:ab OR Guyana:ab OR Suriname:ab OR Bahamas:ab OR Belize:ab OR Aruba:ab or Guadeloupe:ab OR Martinique:ab OR Barbados:ab OR Curacao:ab OR Curaçao:ab OR 'Saint Lucia':ab OR Grenada:ab OR Aruba:ab OR 'Saint Vincent':ab) AND 'the Grenadines':ab OR 'United States Virgin Islands':ab OR Antigua:ab) AND Barbuda:ab OR Dominica:ab OR 'Cayman Islands':ab OR Ssaint Kitts':ab or “British Guiana*” or “French West Indies”) AND Nevis:ab OR Turks:ab) AND 'Caicos Islands':ab) OR 'Sint Maarten':ab OR 'Saint Martin':ab OR 'British Virgin Islands':ab OR ‘Saint Vincent and the Grenadines':ab or ‘Caribbean Netherlands':ab OR Anguilla:ab OR 'Saint Barthélemy':ab OR 'Saint Barthelemy':ab OR Montserrat:ab or ‘British West Indies’:ab) AND 'Caribbean':ab,ti | |
| **LILACS** | **136** |
| ((leptospirosis ) OR (neglected tropical disease) OR zoonosis AND (haiti OR "Dominican Republic" OR cuba OR "Puerto Rico" OR Jamaica OR "Trinidad and Tobago" OR guyana OR suriname OR bahamas OR belize OR guadeloupe OR martinique OR barbados OR curacao OR curaçao OR "Saint Lucia" OR grenada OR aruba OR "Saint Vincent and the Grenadines" OR "United States Virgin Islands" OR "Antigua and Barbuda" OR dominica OR "Cayman Islands" OR "Saint Kitts and Nevis" OR "Turks and Caicos Islands" OR "Sint Maarten" OR "Saint Martin" OR "British Virgin Islands" OR "Caribbean Netherlands" OR anguilla OR "Saint Barthélemy" OR "Saint Barthelemy" OR Montserrat or “British Guiana” or “French West Indies” or “British West Indies”) OR (caribbean) AND (patients) OR (humen)) AND instance:"lilacsplus" AND instance:"lilacsplus" AND instance:"lilacsplus" AND instance:"lilacsplus" AND instance:"lilacsplus" AND instance:"lilacsplus" AND instance:"lilacsplus" | |
