## Supplementary material for "Leptospirosis in the Caribbean Region between 2000 and 2022 – a Scoping Review of Morbidity and Mortality": S3 Table

**Supporting Table 3. The complete list of the 48 publications (no time restriction) reporting leptospirosis cases or seroprevalence in at least one of the 27 CRICTs and the period in which the study was conducted.**

| **First author, year** | **Study years** | **Anguilla** | **Antigua and Barbuda** | **Barbados** | **British Virgin Islands** | **Cayman Islands** | **Cuba** | **Dominica** | **Dominican Republic** | **Grenada** | **Guadeloupe** | **Haiti** | **Jamaica** | **Martinique** | **Montserrat** | **Puerto Rico** | **Saint Kitts and Nevis** | **Saint Lucia** | **Saint Vincent and the Grenadines** | **Trinidad and Tobago** | **U.S. Virgin Islands** |
| --- | --- | --- | --- | --- | --- | --- | --- | --- | --- | --- | --- | --- | --- | --- | --- | --- | --- | --- | --- | --- | --- |
| Adesiyun 2010 | 2006 |  |  |  |  |  |  |  |  |  |  |  |  |  |  |  |  |  |  | x |  |
| Adesiyun, 2011 | 1997-2005 | x | x |  | x | x |  | x |  | x |  |  | x |  |  |  | x | x | x | x |  |
| Artus 2022 | 2019 |  |  |  |  |  |  |  |  |  |  |  |  |  |  |  |  |  |  |  | x |
| Batchelor, 2012 | 1992-2007 |  |  |  |  |  |  |  |  |  |  |  |  |  |  |  |  |  |  |  |  |
| Bennet, 1991 | 1979-1986 |  |  | x |  |  |  |  |  |  |  |  |  |  |  |  |  |  |  |  |  |
| Briskin 2019 | 2015 |  |  |  |  |  |  |  |  |  |  |  |  |  |  | x |  |  |  |  |  |
| Bruce 2005 | 1996-1997 |  |  |  |  |  |  |  |  |  |  |  |  |  |  | x |  |  |  |  |  |
| Cassadou, 2016 | 2011 |  |  |  |  |  |  |  |  |  | x |  |  | x |  |  |  |  |  |  |  |
| Chery, 2020 | 2010-2017 |  |  |  |  |  |  |  |  |  |  |  |  |  |  |  |  | x |  |  |  |
| Damude, 1979*^1^ | 1968-1974 |  |  | x |  |  |  |  |  |  |  |  |  |  |  |  |  |  |  |  |  |
| Damude, 1979*^2a^ | 1975-1977 |  |  | x |  |  |  |  |  |  |  |  |  |  |  |  |  |  |  |  |  |
| Damude, 1979*2^b^ | 1975-1977 |  |  | x |  |  |  |  |  |  |  |  |  |  |  |  |  |  |  |  |  |
| Damude, 1979*2^c^ | 1975-1977 |  |  | x |  |  |  |  |  |  |  |  |  |  |  |  |  |  |  |  |  |
| Edwards, 1990 | 1983-1986 |  |  | x |  |  |  |  |  |  |  |  |  |  |  |  |  |  |  |  |  |
| Everard 1985 | 1977-1978 |  |  |  |  |  |  |  |  |  |  |  |  |  |  |  |  |  |  | x |  |
| Everard 1987 | 1977-1982 |  |  |  |  |  |  |  |  |  |  |  |  |  |  |  |  |  |  | x |  |
| Everard 1989 | 1980-1983 |  |  |  |  |  |  |  |  |  |  |  |  |  |  |  |  |  |  | x |  |
| Everard, 1979a*1 | 1975-1978 |  |  |  |  |  |  |  |  | x |  |  |  |  |  |  |  |  |  |  |  |
| Everard, 1979a*2 | 1975-1978 |  |  |  |  |  |  |  |  | x |  |  |  |  |  |  |  |  |  |  |  |
| Everard, 1984 | 1979-1982 |  |  | x |  |  |  |  |  |  |  |  |  |  |  |  |  |  |  |  |  |
| Everard, 1992 | 1979-1982 |  |  | x |  |  |  |  |  |  |  |  |  |  |  |  |  |  |  |  |  |
| Everard, 1995 | 1979-1991 |  |  | x |  |  |  |  |  |  |  |  |  |  |  |  |  |  |  |  |  |
| Gale, 1990 | 1984-1988 |  |  | x |  |  |  |  |  |  |  |  |  |  |  |  |  |  |  |  |  |
| Gentilini, 1964*1 | 1959 |  |  |  |  |  |  |  |  |  |  | x |  |  |  |  |  |  |  |  |  |
| Gentilini, 1964*2 | 1959 |  |  |  |  |  |  |  |  |  |  | x |  |  |  |  |  |  |  |  |  |
| Golden, 2014 | 2007-2008 |  |  |  |  |  | x |  |  |  |  |  |  |  |  |  |  |  |  |  |  |
| Gonzalez, 1976 | 1973 |  |  |  |  |  | x |  |  |  |  |  |  |  |  |  |  |  |  |  |  |
| Grant, 1964*1 | 1953-1963 |  |  |  |  |  |  |  |  |  |  |  | x |  |  |  |  |  |  |  |  |
| Grant, 1964*2 | 1953-1963 |  |  |  |  |  |  |  |  |  |  |  | x |  |  |  |  |  |  |  |  |
| Herman-Storck, 2005 | 1994-2001 |  |  |  |  |  |  |  |  |  | x |  |  |  |  |  |  |  |  |  |  |
| Herman-Storck, 2008 | 2003-2004 |  |  |  |  |  |  |  |  |  | x |  |  |  |  |  |  |  |  |  |  |
| Hiatt 1976 | 1974 |  |  |  |  |  |  |  |  |  |  |  |  |  |  | x |  |  |  |  |  |
| James 2013 | 2010-2011 |  |  |  |  |  |  |  |  |  |  |  |  |  |  |  |  |  |  | x |  |
| Jones 2024 | 2022 |  |  |  |  |  |  |  |  |  |  |  |  |  |  | x |  |  |  |  |  |
| Levett, 2000 | 1995-1997 |  |  | x |  |  |  |  |  |  |  |  |  |  |  |  |  |  |  |  |  |
| Lhomme, 1996 | 1987-1992 |  |  |  |  |  |  |  |  |  |  |  |  | x |  |  |  |  |  |  |  |
| Lindo, 2013 | 2007-2008 |  |  |  |  |  |  |  |  |  |  |  | x |  |  |  |  |  |  |  |  |
| Mohan 2009 | 1996-2007 |  |  |  |  |  |  |  |  |  |  |  |  |  |  |  |  |  |  | x |  |
| Nilles, 2024 | 2021 |  |  |  |  |  |  |  | x |  |  |  |  |  |  |  |  |  |  |  |  |
| Perez, 1998 | 1987-1993 |  |  |  |  |  | x |  |  |  |  |  |  |  |  |  |  |  |  |  |  |
| Sanchez, 1993 | 1986-1990 |  |  |  |  |  | x |  |  |  |  |  |  |  |  |  |  |  |  |  |  |
| Sanders 1999 | 1996 |  |  |  |  |  |  |  |  |  |  |  |  |  |  | x |  |  |  |  |  |
| Sharp 2016 | 2010 |  |  |  |  |  |  |  |  |  |  |  |  |  |  | x |  |  |  |  |  |
| Strobel, 1992 | 1989 |  |  |  |  |  |  |  |  |  | x |  |  |  |  |  |  |  |  |  |  |
| Suarez-Hernandez, 1999 | 1982-1995 |  |  |  |  |  | x |  |  |  |  |  |  |  |  |  |  |  |  |  |  |
| Suarez-Hernandez, 2001 | 1998 |  |  |  |  |  | x |  |  |  |  |  |  |  |  |  |  |  |  |  |  |
| Suarez-Hernandez. 1995 | 1984-1988 |  |  |  |  |  | x |  |  |  |  |  |  |  |  |  |  |  |  |  |  |
| Villafranca, 2002 | 1996-1998 |  |  |  |  |  | x |  |  |  |  |  |  |  |  |  |  |  |  |  |  |
| Wood, 2014 | 2009-2011 |  | x |  |  |  |  | x |  | x |  |  | x |  | x |  | x | x | x |  |  |
