## Supplementary material for "Leptospirosis in the Caribbean Region between 2000 and 2022 – a Scoping Review of Morbidity and Mortality": S4 Table

**Supporting Table 4. The complete list of the 16 publications reporting leptospirosis cases or seroprevalence in at least one of the 27 CRICTs after the year 2000 included in the scoping review.**

| First author, year | Year | | | | | | | | | | | | | | | | | | | | | | |
| --- | --- | --- | --- | --- | --- | --- | --- | --- | --- | --- | --- | --- | --- | --- | --- | --- | --- | --- | --- | --- | --- | --- | --- |
|  | 2000 | 2001 | 2002 | 2003 | 2004 | 2005 | 2006 | 2007 | 2008 | 2009 | 2010 | 2011 | 2012 | 2013 | 2014 | 2015 | 2016 | 2017 | 2018 | 2019 | 2020 | 2021 | 2022 |
| Wood, 2014 |  |  |  |  |  |  |  |  |  | x | x | x |  |  |  |  |  |  |  |  |  |  |  |
| Golden, 2014 |  |  |  |  |  |  |  | x | x |  |  |  |  |  |  |  |  |  |  |  |  |  |  |
| Nilles, 2024 |  |  |  |  |  |  |  |  |  |  |  |  |  |  |  |  |  |  |  |  |  | x |  |
| Herrmann-Storck, 2005 | x |  |  |  |  |  |  |  |  |  |  |  |  |  |  |  |  |  |  |  |  |  |  |
| Herrmann-Storck, 2008 |  |  |  | x | x |  |  |  |  |  |  |  |  |  |  |  |  |  |  |  |  |  |  |
| Cassadou, 2016 |  |  |  |  |  |  |  |  |  |  |  | x |  |  |  |  |  |  |  |  |  |  |  |
| Chery, 2020 |  |  |  |  |  |  |  |  |  |  | X | X | X | X | X | X | X | x |  |  |  |  |  |
| Batchelor, 2012 | x | x | x | x | x | x | x | x |  |  |  |  |  |  |  |  |  |  |  |  |  |  |  |
| Lindo, 2013 |  |  |  |  |  |  |  | x | x |  |  |  |  |  |  |  |  |  |  |  |  |  |  |
| Mohan, 2009 | x | x | x | x | x | x | x | x |  |  |  |  |  |  |  |  |  |  |  |  |  |  |  |
| James, 2013 |  |  |  |  |  |  |  |  |  |  | x | x |  |  |  |  |  |  |  |  |  |  |  |
| Adesiyun, 2010 |  |  |  |  |  |  | x |  |  |  |  |  |  |  |  |  |  |  |  |  |  |  |  |
| Artus, 2022 |  |  |  |  |  |  |  |  |  |  |  |  |  |  |  |  |  |  |  | x |  |  |  |
| Sharp, 2016 |  |  |  |  |  |  |  |  |  |  | x |  |  |  |  |  |  |  |  |  |  |  |  |
| Briskin, 2019 |  |  |  |  |  |  |  |  |  |  |  |  |  |  |  | x |  |  |  |  |  |  |  |
| Jones, 2024 |  |  |  |  |  |  |  |  |  |  |  |  |  |  |  |  |  |  |  |  |  |  | x |
