## Supplementary material for "Leptospirosis in the Caribbean Region between 2000 and 2022 – a Scoping Review of Morbidity and Mortality": S5 Table

**Supporting Table** **5. Summary of characteristics of publications based on routine surveillance-based studies.**

| First author, year | Study aims | | | | | | Study design | Diagnostic test | |
| --- | --- | --- | --- | --- | --- | --- | --- | --- | --- |
|  | Motivated by an extreme weather event? | Estimate Incidence | Identify risk factors | Lab confirmed/ suspected cases | Impact of PH intervention | Describe changes through time | Reporting Site | ELISA | MAT |
| Golden, 2014 | Yes |  |  |  | x |  | Mixed | NR | NR |
| Herman-Storck, 2008 | No | x | x |  |  | x | Hospital | X | X |
| Cassadou, 2015 | No | x |  |  |  |  | Mixed | X | X |
| Batchelor, 2012 | No | x | x |  |  |  | Mixed | X | X |
| Mohan, 2009 | No | x | x |  |  | x | Mixed | X |  |
| Sharp, 2016 | No |  |  | x |  |  | Mixed | X | X |
| Jones, 2024 | Yes |  | x | x |  |  | Mixed | X |  |
| Herman-Storck, 2005 | No |  | X | X |  |  | Hospital |  | X |
| Chery et al, 2020 | Yes | X | X | X |  |  | Mixed | X |  |
