## Supplementary material for "Leptospirosis in the Caribbean Region between 2000 and 2022 – a Scoping Review of Morbidity and Mortality": S6 Table

**Supporting Table 6. Summary of characteristics of publications based on seroprevalence studies.**

| First author, year | Study aims | | | | Study design | | | Diagnostic test | |
| --- | --- | --- | --- | --- | --- | --- | --- | --- | --- |
|  | Motivated by an extreme weather event? | Identify risk factors | Risk group prevalence | Lab confirmed/ suspected cases | Sampling design | Recruitment site | Household setting | ELISA | MAT |
| Wood, 2014 | No |  | x |  | NR | NR | NR | X |  |
| Nilles, 2021 | No | x |  |  | Three-stage, hierarchical random sampling | Community | Mixed |  | X |
| Lindo, 2013 | No |  |  | x | Random | Mixed | Mixed | X |  |
| James, 2013 | No |  | x |  | Convenience | Community | Mixed | X | X |
| Adesiyun, 2010 | No |  | x |  | Convenience | Community | Rural | X |  |
| Artus, 2022 | Yes | x |  |  | A stratified, random two-stage cluster sampling design | Community | Mixed |  | X |
| Briskin, 2019 | Yes | x |  |  | Convenience | Community | Urban |  | X |
