## Supplementary material for "Leptospirosis in the Caribbean Region between 2000 and 2022 – a Scoping Review of Morbidity and Mortality": S7 Table

**Supporting Table 7. Case definition and laboratory tests used to confirm cases presented in routine surveillance-based studies.**

| First author, year | Case definition |
| --- | --- |
| Sharp, 2016 | Confirmed: Antigen positive by IHC, or PCR positive, or fourfold increase in the MAT between two paired samples (usually acute phase and convalescent phase), one isolated MAT sample > 1:800 or ELISA IgM positive  Probable: MAT<800. |
| Jones, 2013 | Confirmed: PCR positive and Probable ELISA IgM positive (MAT was not possible during the study) |
| Herrmann-Storck, 2005 | Confirmed: ELISA IgM >1:400, fourfold increase between two MAT samples (acute and convalescent phase).  Suspected: ELISA IgM between 100 and 400, or an increase on the MAT results between paired samples less than fourfold. |
| Chery, 2020 | ELISA IgM >1:400. |
| Mohan, 2009 | Confirmed: ELISA IgM > 1:640  Probable: ELISA IgM between 1:80 and 1:320 |
| Batchelor, 2012 | MAT 1:200  ELISA IgM > 1:320 |
| Cassadou, 2016 | Suspected cases were investigated with PCR, if positive, case was confirmed. If negative, a ELISA IgM was performed. If positive (>1:400), a MAT was performed. MAT were considered positive if >1:400. |
| Herrmann-Storck, 2008 | fourfold increase between two MAT samples (acute and convalescent phase).  ELISA IgM > 1:400 |
| Golden, 2014 | No case definition criteria presented. |
