## Supplementary material for "Leptospirosis in the Caribbean Region between 2000 and 2022 – a Scoping Review of Morbidity and Mortality": S8 Table

**Supporting Table 8. Laboratory test used to identify seropositive individuals in seroprevalence studies**

| First author, year | Laboratory test |
| --- | --- |
| Lindo, 2013 | ELISA IgM |
| Wood, 2014 | ELISA IgG |
| Nilles, 2024 | MAT > 1:100 |
| James, 2013 | ELISA IgG and MAT > 1:20 |
| Adesiyun, 2010 | ELISA IgM 1:60 |
| Artus, 2020 | MAT > 1:100 |
| Briskin, 2019 | MAT 1:50 |
